# Auditory Profile-based Hearing-aid Fitting: A Proof-Of-Concept Study

**DOI:** 10.1101/2020.04.14.20036459

**Authors:** Raul Sanchez-Lopez, Michal Fereczkowski, Sébastien Santurette, Torsten Dau, Tobias Neher

## Abstract

**Objective:** The clinical characterization of hearing deficits for hearing-aid fitting purposes is typically based on the pure-tone audiogram only. In a previous study, a group of hearing-impaired listeners were tested using a comprehensive test battery designed to tap into different aspects of hearing. A data-driven analysis of the data yielded four clinically relevant patient subpopulations or “auditory profiles”. In the current study, profile-based hearing-aid settings were proposed and evaluated to explore their potential for providing more targeted hearing-aid treatment.

**Design:** Four candidate hearing-aid settings were implemented and evaluated by a subset of the participants tested previously. The evaluation consisted of multi-comparison preference ratings carried out in realistic sound scenarios.

**Results:** Listeners belonging to the different auditory profiles showed different patterns of preference for the tested hearing-aid settings that were largely consistent with the expectations.

**Conclusion:** The results of this proof-of-concept study support further investigations into stratified, profile-based hearing-aid fitting with wearable hearing aids.

## Introduction

Hearing loss (HL) is typically treated with hearing aids (HA). The primary purpose of HAs is to provide gain to the input signal to compensate for reduced audibility. In addition, modern HA incorporate advanced signal processing algorithms for noise suppression (Chung 2004). As a consequence, numerous parameters need to be adjusted as part of the hearing-aid fitting process.

In current clinical practice, the assessment of the hearing deficits of a patient relies mainly on pure-tone audiometry. Based on a fitting rule that typically only uses the audiogram of the patient as input, the HA amplification is then adjusted. For example, the “National Acoustic Laboratories – Nonlinear 2” fitting rule (Keidser et al. 2011) is commonly used for this. The NAL-NL2 rule relies on a combination of empirical knowledge and modelling aimed at maximizing the effective speech audibility. Even though this can provide a reasonable overall solution, there are also patients whose hearing difficulties are not captured by the audiogram and who may therefore benefit from other fitting strategies (Keidser & Grant 2001; Oetting et al. 2018; Henry et al. 2019). Such fitting strategies could include the adjustment of advanced HA features, which are not yet incorporated into existing fitting rules. For example, noise reduction and directional processing are currently activated based on lifestyle considerations rather than audiological factors. Although advanced HA features can improve the signal-to-noise ratio (SNR), preference for these settings can vary substantially across listeners, possibly because of unwanted speech distortions that these algorithms typically also introduce (Neher et al. 2016). Therefore, it is possible that the individualized adjustment of speech enhancement algorithms could improve HA outcome, for example for patients with poor speech intelligibility in challenging environments.

In a recent study, we identified four clinically relevant subgroups of hearing-impaired (HI) listeners using a data-driven approach (Sanchez-Lopez et al. 2020a). The listeners were characterized by their degree of perceptual deficits or “distortions”, which were estimated using a battery of auditory tests tapping into loudness and speech perception, binaural processing abilities and spectro-temporal resolution (Sanchez-Lopez et al. 2020b). Four archetypal patterns of perceptual deficits – referred to as Profiles A, B, C and D – were uncovered. These profiles varied along two primary dimensions, or types, of deficits: speech intelligibility (SI) and loudness perception (LP) related deficits. Overall, this suggested that these profiles could benefit from more tailored HA solutions.

In the medical field, personalized treatment aims at providing tailored solutions to clinically relevant subgroups of patients. Here, a number of profile-based candidate hearing-aid settings (HAS) were evaluated. Listeners with a high degree of LP-related deficits (Profiles C and D) were expected to prefer a gain prescription aimed at loudness normalization (Oetting et al. 2018), whereas listeners with a high degree of SI-related deficits (Profiles B and C) were expected to prefer HAS with advanced signal processing (**Figure S 1**). As such, the present study examined the validity of auditory profile-based HA fitting with respect to the listeners’ subjective preference. A multi-comparison sensory evaluation (Zacharov 2018) was performed with a group of participants who had previously been classified into the four auditory profiles. This made it possible to explore whether listeners belonging to different auditory profiles would exhibit different patterns of HA outcome.

## Results and discussion

Four candidate HAS (HAS-I, HAS-II, HAS-III and HAS-IV) were evaluated together with a standard clinical HAS (HAS-O). In HAS-I and HAS-II, fast-acting compression was applied to provide non-linear gain according to an audibility-based prescription formula. In HAS-III and HAS-IV, slow-acting compression based on the principle of loudness normalization was applied. Furthermore, in HAS-II and HAS-III advanced HA features were activated to provide about 2.5 dB of SNR improvement under noisy conditions (see Method, Error! Reference source not found., Error! Reference source not found. for more details).

Figure 1 shows the mean preference ratings for profiles A, B, C and D under *quiet* conditions. Profile-A listeners preferred HAS-O over the two HAS with fast-acting compression across the tested presentation levels (55, 65 and 75 dB sound pressure level, SPL). Profile-B listeners preferred HAS-I over HAS-III at 65 and 75 dB SPL; at 55 dB SPL they provided the highest rating to HAS-IV. Profile-C and -D listeners showed a preference for HAS-IV and consistently disliked HAS-I.

**Figure 1:**
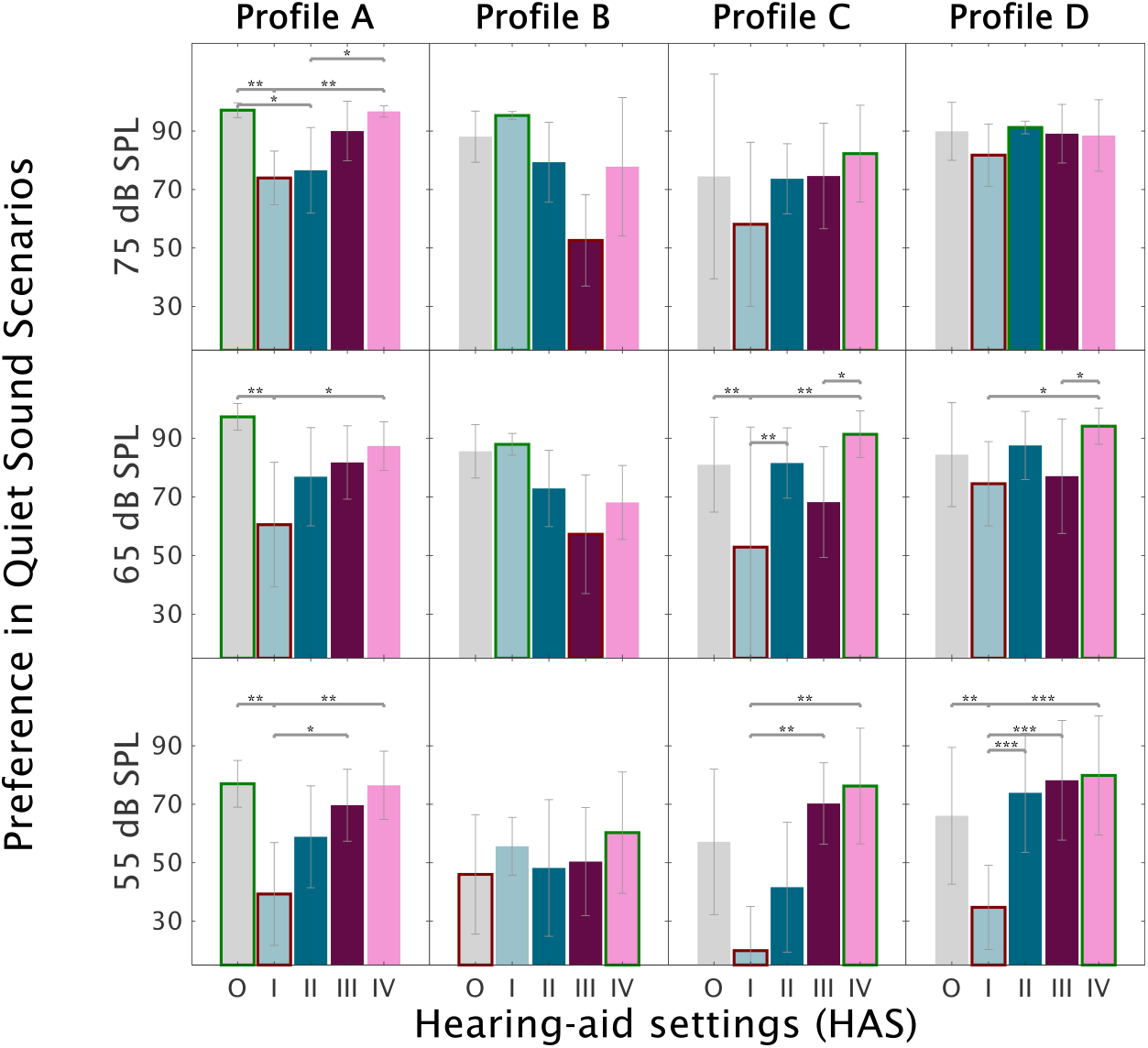
Mean preference ratings for the evaluated HAS (O-IV) under *quiet* conditions across three level conditions: 55 dB SPL (bottom panels), 65 dB SPL (middle panels) and 75 dB SPL (top panels). The highest (best) ratings are highlighted by green borders and the lowest by red borders. The columns represent the results of the listeners belonging to profile A (left), B (mid-left), C (mid-right) and D (right). Error bars show ±1 standard deviation. Signficant differences according to a two-way ANOVA with repetition and participant as factors followed by Tukey’s honest significant differences tests are marked by asterisks. (***) *p*<0.001, (**) *p*<0.01, (*) *p*<0.05.

Figure 2 shows the mean preference ratings under *noisy* conditions. Profile-A listeners preferred HAS-III and HAS-O over HAS-I. Profile-B listeners consistently disliked HAS-III and showed a preference for HAS-O, HAS-I and HAS-II. Profile-C listeners preferred HAS-III over the other HAS at higher SNRs (0 and +4 dB). However, HAS-O was also preferred at lower SNRs. Profile-D listeners showed only significant differences at +4 dB SNR, with HAS-IV receiving the highest ratings and HAS-I the lowest ratings.

**Figure 2:**
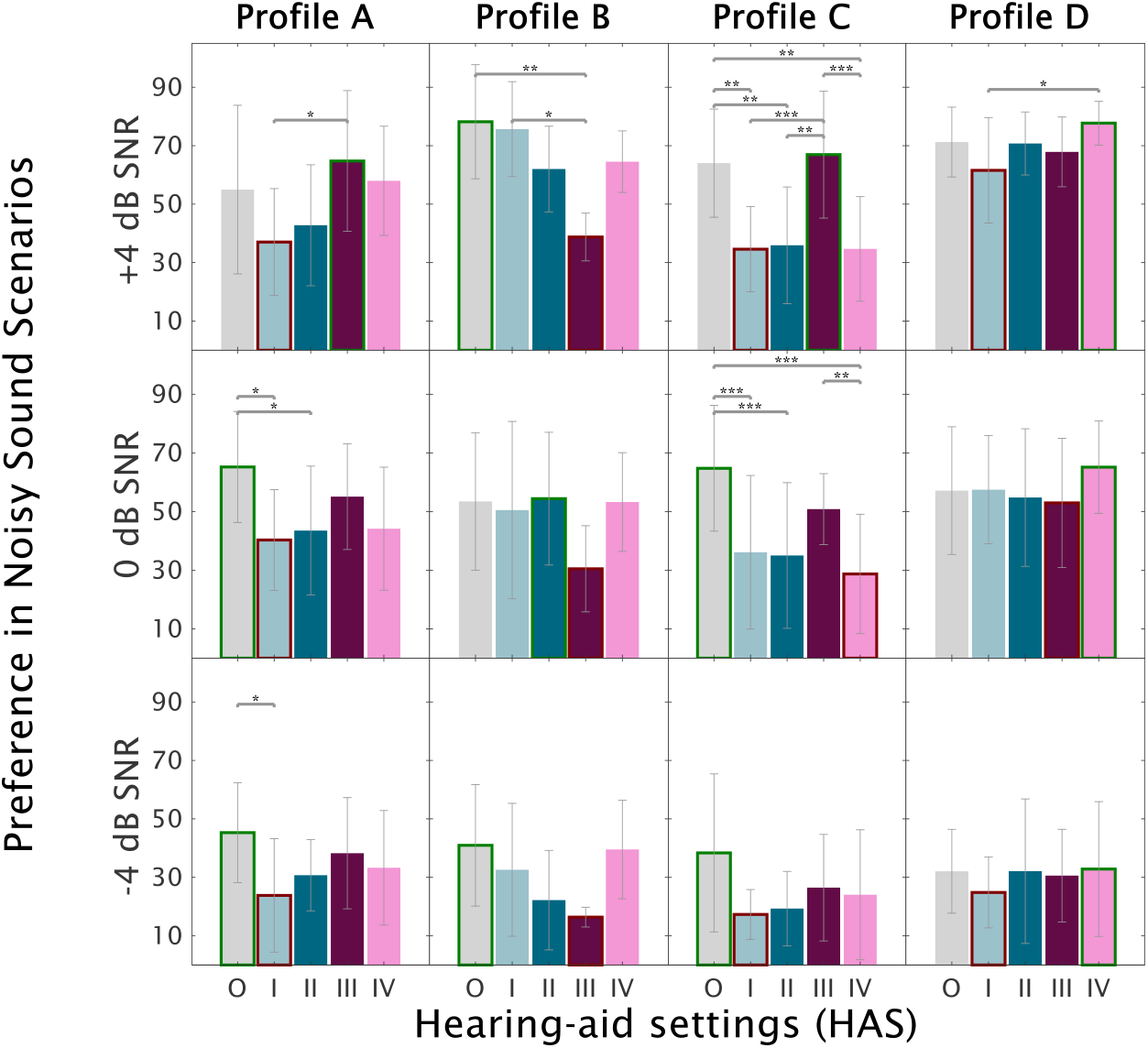
Mean preference ratings for the evaluated HAS (O-IV) under *noisy* conditions across the three SNR conditions: -4 dB SNR (bottom panels), 0 dB SNR (middle panels) and +4 dB SNR (top panels). The highest (best) ratings are highlighted by green borders and the lowest by red borders. The columns represent the results of the listeners belonging to profile A (left), B (mid-left), C (mid-right) and D (right). Error bars show ±1 standard deviation. Signficant differences according to a three-way ANOVA with repetition, noise type and participant as factors followed by Tukey’s honest significant differences tests are marked by asterisks. (***) *p*<0.001, (**) *p*<0.01, (*) *p*<0.05.

The current study aimed to identify patterns of HAS preference in listeners belonging to four distinct auditory profiles. The obtained results suggest that Profile-A and -C listeners based their judgements on similar criteria, especially under noisy conditions. In contrast, Profile-B and -D listeners showed significantly different patterns. While Profile-B listeners disliked the HAS with loudness-based gain prescription and SNR improvement, Profile-D listeners favored loudness-based gain prescription and showed no preference for SNR improvement. The results obtained for the quiet condition support the use of loudness-based gain prescriptions for profiles with a high degree of LP-related deficits. In contrast, SNR improvement was only preferred by one of the two profiles showing a high degree of SI-related deficits (Profile C) when tested at positive SNRs; it could be that the other profile (B) is more susceptible to, and therefore dislikes more, the distortions introduced by the noise suppression algorithm. However, Profile-B listeners showed a preference for fast-acting compression, which is consistent with previous research findings concerning HA outcome for listeners with steeply sloping hearing losses (Gatehouse et al. 2006).

In summary, Profile-A and -B listeners preferred audibility-based gain prescriptions, whereas Profile-C and -D listeners preferred loudness-based gain prescriptions. Besides, SNR improvement was beneficial for Profile-C listeners (with a high degree of SI-related deficits). Overall, these initial findings provide a useful basis for further investigations into profile-based HA fitting strategies that will include field studies with wearable devices and speech intelligibility asssessments.

## Methods

Seven listeners participated in the current study (N=2 in each subgroup except for Profile-B, N=1). All of them had previously completed a comprehensive auditory test battery (Sanchez-Lopez et al. 2020b), based on which they had been classified as belonging to one of the four auditory profiles (Sanchez-Lopez et al. 2020a). The study was approved by the Science-Ethics Committee for the Capital Region of Denmark H-16036391.

For the study, a hearing-aid simulator (HASIM) was used that consisted of three stages: A beamforming stage, a noise reduction stage and an amplitude compression stage (see **Table S 1** for details). The beamformer and noise reduction settings were selected based on the achievable SNR improvement (Sanchez-Lopez et al. 2018).

Nine sound scenarios were tested. In each scenario, a fragment of a realistic conversation taken from a publicly available database (Sørensen et al. 2018) was used for engaging the listener in the sound scene. The participant was instructed to listen actively to the conversation. The tested sound scenarios differed in terms of the background noise. Three noise conditions were included: 1) *cafeteria* noise (presentation level of 65 dB SPL), 2) *traffic* noise (presentation level of 75 dB SPL), and 3) *quiet*. Furthermore, there were three SNR or level conditions. That is, in the case of the *cafeteria* and *traffic* scenarios the target was scaled in level to achieve SNRs of -4, 0 or +4 dB. In the *quiet* scenario, the target presentation level was either 55, 65 or 75 dB SPL.

The multi-comparison of the HAS was realized using the SenselabOnline software (SenseLab dept. 2017). On a given trial, six stimuli were presented to the listener: An anchor resembling a ‘broken’ hearing aid, a clinically representative HAS, and the four candidate HAS (I, II, III and IV). The multi-comparisons were performed sequentially across several trials. In each case, a 20-sec audio file corresponding to a given sound scenario processed with the HASIM was played back (**Figure S 2**). The listeners then used a slider ranging from 0 to 100 to rate the sound of each HAS. The question posed to the listeners was “Which hearing aid would you choose?”. When giving their ratings, they were instructed to focus on overall preference rather than on specific attributes such as noise annoyance or speech clarity.

## Data Availability

The materials and data are available in a Zenodo Repository DOI:10.5281/zenodo.3732320

https://zenodo.org/record/3732320

## Acknowledgement

The authors thank SenseLab (Force technology) and S.G. Nielsen for their input and support and the listeners for their participation. This work was supported by Innovation Fund Denmark Grand Solutions 5164-00011B (Better hEAring Rehabilitation project) and the other BEAR partners. The funding and collaboration of all partners is sincerely acknowledged.

